# A comparative study of multidrug resistant bacteria (MDRB) isolated from various patients in different wards of a hospital during post Covid-19

**DOI:** 10.1101/2024.10.25.24315976

**Authors:** Abhishek Ojha, Dwight Figueiredo, Shabnum Khan

**Author notes:** Corresponding author; Abhishek Ojha.

## Abstract

Antibacterial resistance is an alarming global concern and a public health challenge of the twenty-first century for which effective systems are required to track and treat ABR. We performed microbiological and antibiotic susceptibility testing on the samples to detect and characterize Multidrug-Resistant Bacteria (MDRB) isolated from patients segregating MDRB characteristics (types, prevalence and distribution of MDRB) based on time (i.e., during versus post covid-19) and location (i.e., different wards of a tertiary care hospital). We observed an increase of MDRB in 2022 as compared to 2021 and 2023. These MDRB had a Shannon and Simpson index values of 1.138 to 1.508 and 0.643 to 0.775, respectively and an observed evenness values of 0.780 to 1.042, which revealed the microbial diversity recovered from the patient samples. In keeping with previous MDR studies, *Klebsiella*, *E*. *coli*, *Citrobacter*, *Acinetobacter* and *Pseudomonas* were identified from the patient samples. Moreover, compared to previous reports, the percentage of MDR-bacteria, i.e., *Klebsiella* (40 %), *E*. *coli* (28 %), and *Citrobacter* (19 %), populations were higher in this study. We observed that Gamma-proteobacteria were predominant across all the recovered samples, and that *Acinetobacter* and *Klebsiella* isolated from the samples were 100 % resistant to twenty and eleven antibiotics, respectively. Furthermore, the fatality rate was low compared with the available reports suggesting possibilities for effective recovery if given rapid and tailored treatment. Given the challenges faced by MDRB strains more surveillance and tracking is needed to ensure effective and specifically targeted treatment strategies.

## 1. Introduction

Over the previous decades, it has been observed that antibiotic resistance is increasing to precariously high levels all over the world as new mechanisms of resistance are looming and spreading worldwide (Alós et al., 2015; Hernando-Amado et al., 2019). Widespread, non-specific, and uninterrupted use of antibacterial antibiotics in treating bacterial infections has been well documented in fuelling resistance among the distinct elements of bacterial populations (Klein et al., 2018; Carvalho et al., 2022). Due to the continuous need of antibacterial substances active against resistant gram-negative microflora, gram-positive multi drug resistant (MDR) pathogens have been overwhelmed by gram-negative bacterial infections (Jones, 2001). Among gram-negative microflora, the commonest MDR microbes identified in severe patients are *Pseudomonas aeruginosa*, *Stenotrophomonas maltophilia*, *Acinetobacter* sp., *and Enterobacteriaceae*. *Staphylococcus aureus* (methicillin-resistant) and *Enterococci* sp. (vancomycin-resistant) are the most common gram-positive isolated, although their occurrence is falling (Jones, 2001; Boucher et al., 2009). Hospital-acquired infections (HAIs) have been known for over a century as a critical clinical issue affecting healthcare quality, and they are the primary source of unfavourable healthcare outcomes (Aly et al., 2008; Memish and El-Saed, 2009; Nannini et al., 2009). The development of MDR microbes (MDRM) in the patient has become a public health issue, emerging as a new concern in many parts of the health system or hospitals (Jindal et al., 2015; Dogru et al., 2010; Teng et al., 2009; Aly et al., 2008). There is extensive use of antibiotics as a drug in critical care units, which establishes a selection burden and stimulates the development of MDRM (Teng et al., 2009; Aly et al., 2008). Further, an intensive care unit (ICU) patient is known to have a high risk of infection due to their underlying health conditions, exposure to various invasive devices, and weakened immunity (Dettenkofer et al., 2001; Ylipalosaari et al., 2006). The HAI rate in general wards is lower than that of ICU-HAI (Weinstein, 1998). The ICU-HAI has been associated with higher costs, morbidity, and mortality (Iskandar et al., 2021; Neidell et al., 2012; Montassier et al., 2013; Gastmeier et al., 2005). The objective of this research was to investigate HAI in various (ICUs, neonatal ICU (NICU), outpatient departments (OPD), male wards, female wards, pediatric wards, and medicine wards) departments to reveal the MDR microbial community, the anti-microbial resistance descriptions, including their effect on MDRM-related mortality and comorbidity.

## 2. Materials and methods

### 2.1. Site of study and patients

The study was performed as single-centre retrospective research at a 900-bed hospital (the Symbiosis University Hospital and Research Centre, SUHRC, Pune, Maharashtra, India), providing quality healthcare to the Pune’s surrounding rural and upcoming urban areas. High patient numbers ensure extensive contact with the local community flows daily due to subsidized healthcare facilities and treatment strategies provided. The hospital provides for both inpatient and outpatient patient treatment services. The hospital possesses operation theatres, ICUs, NICUs, Wards, OPDs. During Covid-19 (Coronavirus disease-2019), the hospital ran with specialized wards assigned to Covid-19-positive patients. The standard infection control measures (PPE, personal protective equipment) and sterilization protocols (https://www.england.nhs.uk/national-infection-prevention-and-control-manual-nipcm-for-england/chapter-1-standard-infection-control-precautions-sicps/) were followed to prevent cross-contamination and spread of Covid-19.

### 2.2. Clinical data

A descriptive retrospective investigation of the occurrence of MDR bacteria in patients treated at the hospital from January 2021 to April 2023, patients’ samples were collected from the different hospital departments as a part of routine testing or when infections were suspected /presumed to be the cause underlying diseases (ICU, NICU, OPD, male ward, female ward, paediatric ward, and medicine ward). Clinical data information relevant to this study was collected and extracted from the departments mentioned above, patient medical charts, infection control surveillance forms, and microbiology laboratory results. The clinical data extracted also included - patient symptoms and comorbidities, neoplasm-related data and its treatment, and most importantly, MDR related infectious microorganisms and their resistance pattern (based on sensitivity to antibiotics, which are recorded data from microbiology laboratory culture test blood culture, etc.) MDRs were determined by applying the Centres for Disease Control and Prevention (CDC) parameters (Horan et al., 2008).

### 2.3. Bacterial identification and susceptibility testing

Microbial isolates were determined by applying the BD Phoenix Automated Microbiology System (USA) and Kirby-Bauer disk diffusion technique (Clinical and Laboratory Standards Institute, 2006.). The protocol detailed, by Caroll et al. (2006) and Hudzicki (2009), were applied to identification and antimicrobial susceptibility tests. Microorganism cultures were obtained from patient samples from different anatomical locations on the body. We examined body fluids such as pus, urine, swabs (from the foot and near the colon), sputum, stool, BAL (Bronchoalveolar lavage) fluid, and ET (Endotracheal Secretion) secretion to determine the occurrence of antibiotic-resistant microbes or pathogens across the samples. The isolates *Escherichia coli*, *Klebsiella pneumoniae*, *Pseudomonas aeruginosa*, *Acinetobacter* sp., and *Citrobacter* sp., were examined to be MDR. An infection at many sites in the same patient was revealed as distinct infection events, except that an identical microbe was revealed concurrently. The clinical outcomes were evaluated up to hospital discharge or patient death. For the reasons of this study, patient death was determined concerning the HAIs/ or as non- HAIs-associated in agreement with the professional (Medical) who endorsed the patient death, including clinical chart data examined by three of the authors of this study.

### 2.4. Statistical analysis

Descriptive statistics were used for categorised variables and expressed in terms of per cent frequency. The normal distribution and diversity (alpha and beta) index were applied to determine the variations in MDR-microbial populations between MDR patient samples (collected from 2021, 2022, and 2023 at SUHRC, Pune) used in this study.

## 3. Results

### 3.1. Overview of patient infection across all the samples

One hundred patients aged one day to 98 years were included (data not shown). Eighty-five patients (out of 100 patients) were treated successfully and discharged (**Table 1**, data not shown), five patients (out of 100 patients) died (**Table 1**) in the hospital, and there was no clarity in the reporting of 10 patients’ data whether they discharged or died. Fifty-three patients (53 %), 38 patients (38 %), and nine patients (9 %) were male, female, and child (female), respectively (**Table 1, Fig. S1**) with various clinical characteristics (data not shown and **Table 1)**. An overall lowest death rate of males were observed among patients (**Fig. 1A**). Patients with comorbidities or with no comorbidities are shown in Table 1. 18 patients with no co-existing comorbidities were observed (**Table 1** and data not shown). The medical conditions at presentation or during the hospital study included; UTI (urinary tract infection, 35 % patient), sepsis (16 % patient), foot gangrene (3 % patient), respiratory disease (2 % patient), cancer (1 % patient), hepatic disease (1 % patient), hepatic cyst (1 % patient), asthma (1 % patient), IHD (ischemic heart disease, 1 % patient), CKD (chronic kidney disease,1 % patient), septic shock (1 % patient), post-CAR (chimeric antigen receptors, 1 % patient) T-cell therapy, hyperthyroidism (1 % patient), osteoporosis (1 % patient), hemoperitoneum (1 % patient), multiple vessel (1 % patient), HIE (hypoxic-ischemic encephalopathy, 1 % patient), femur fracture (1 % patient), that were simultaneously present in a patient (**Table 1**). **Table 2** revealed the sample type distribution and predominance of MDR-gram-negative bacteria (MGNB). The sample urine from the patient showed the highest number (61) of MGNB (**Table 2**). In comparison the lowest number (2) of MGNB was present in stool samples (**Table 2**). In terms of bacterial species identified in the samples, MDR-*Klebsiella pneumoniae* was predominant in urine samples (**Table 2**).

**Fig 1.**
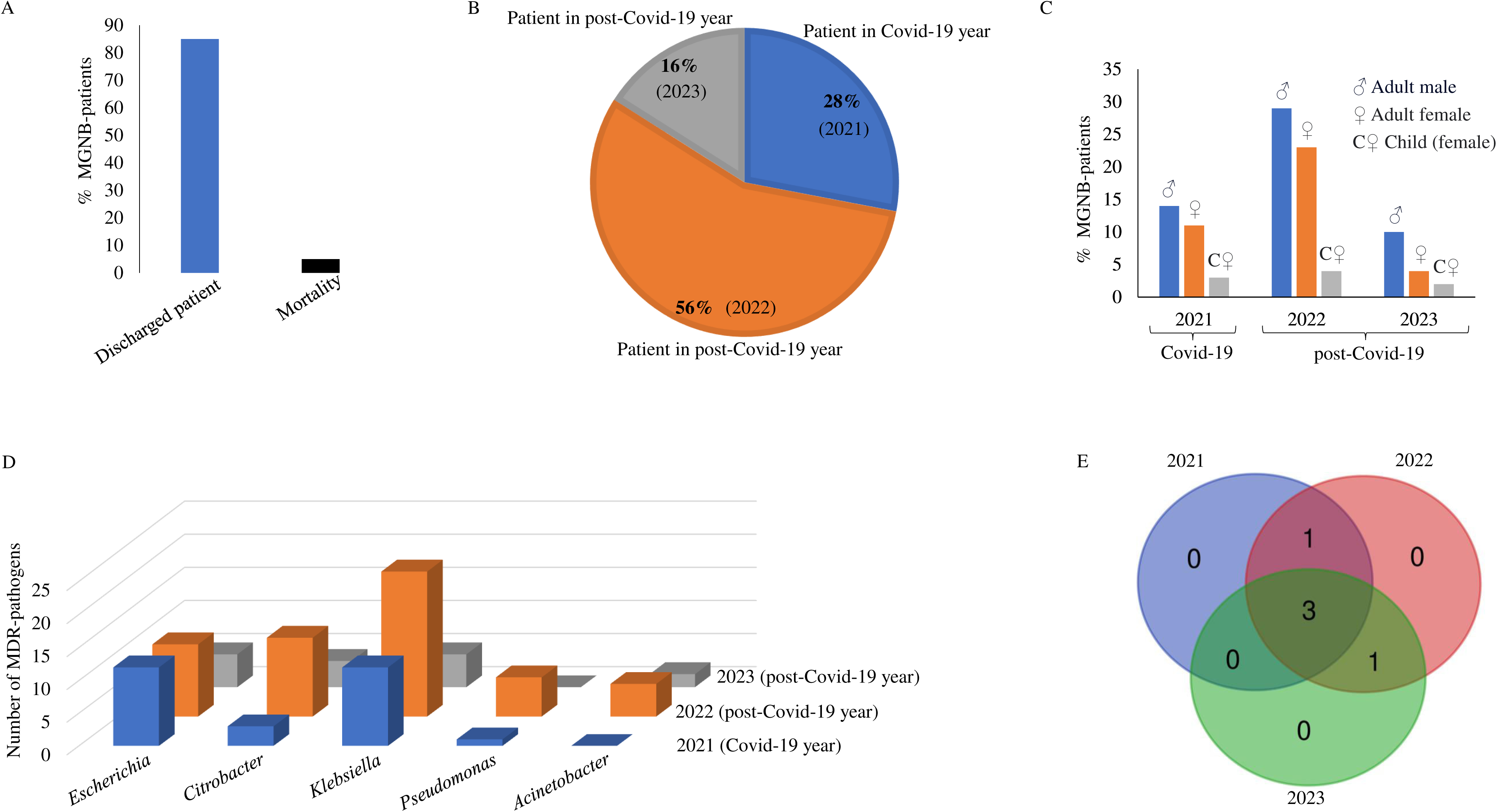
Overview of study. (A) Outcomes of patients with MDR-gram negative bacterial pathogens. (B) Percentage overview of patients with MDR-gram negative bacterial pathogens. (C) Distribution of MDR-gram negative bacterial pathogens in adult male, adult female, and children (female). (D) Relative abundance of different MDR-gram negative bacterial pathogens among all patient samples at the level of genus. (E) Identification of specific and shared MDR-gram negative bacterial pathogens among the patient samples.

**Table 1.** Characteristics of hospital stay of patients with or without comorbidities, 2021-2023 (Covid-19/ or post Covid-19) period.

| S. No. | Characteristic | Patients (N=100) |
| --- | --- | --- |
| 1 | Age (range) | 1 day - 98 years |
| 2 | Discharged | 85 (85 %) |
| 3 | Death | 5 (5 %) |
| 4 | Male (adult) | 53 (53 %) |
| 5 | Female (adult) | 38 (38 %) |
| 6 | Children (female) | 9 (9 %) |
| 7 | Male ward | 27 (27 %) |
| 8 | Female ward | 25 (25 %) |
| 9 | Intensive care unit (ICU) admission | 29 (29 %) |
| 10 | Outpatient department (OPD) | 12 (12 %) |
| 11 | Pediatric ward | 4 (4 %) |
| 12 | Neonatal intensive care unit (NICU) admission | 1 (1 %) |
| 13 | Comorbidities |  |
| a. | no comorbidities | 18 (18 %) |
| b. | Urinary Tract Infection (UTI) | 35 (35 %) |
| c. | Sepsis | 16 (16 %) |
| d. | Foot gangrene | 3 (3 %) |
| e. | Respiratory disease | 2 (1 %) |
| f. | Cancer | 1 (1 %) |
| g. | Hepatic disease | 1 (1 %) |
| h. | Hepatic cyst | 1 (1 %) |
| i. | Asthma | 1 (1 %) |
| j. | Ischemic heart disease (IHD) | 1 (1 %) |
| k. | Chronic kidney disease (CKD) | 1 (1 %) |
| l. | Septic shock | 1 (1 %) |
| m. | Post-compensatory anti-inflammatory response (CAR) | 1 (1 %) |
| n. | Hyperthyroidism | 1 (1 %) |
| o. | Osteoporosis | 1 (1 %) |
| p. | Hemoperitoneum | 1 (1 %) |
| q. | Multiple vessel | 1 (1 %) |
| r. | Hypoxic-ischemic encephalopathy (HIE) | 1 (1 %) |
| s. | Femur fracture | 1 (1 %) |

**Table 2.**
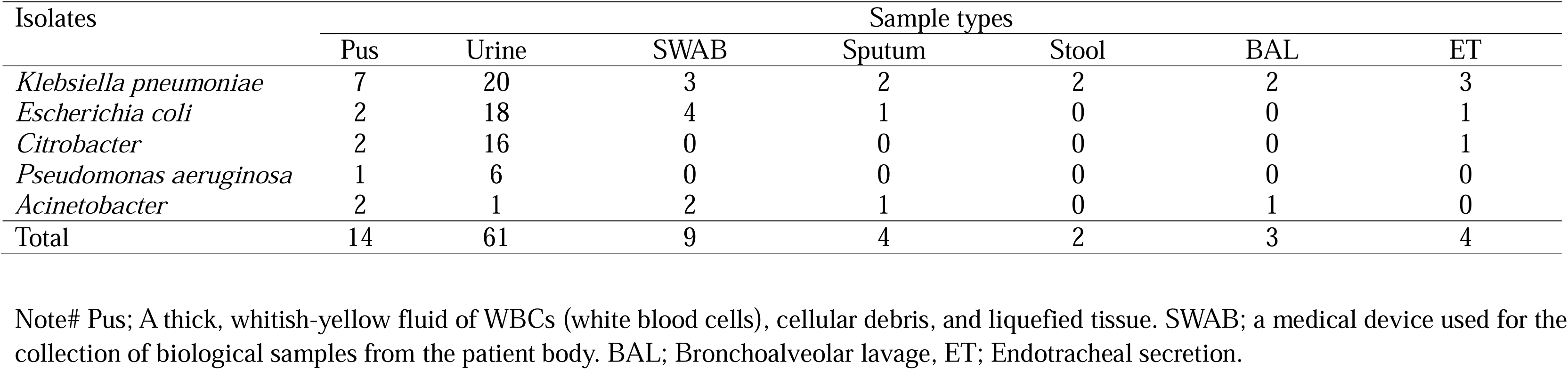
Dominancy of MDR bacterial isolates from different types of patient’s sample.

| Isolates | Sample types |  |  |  |  |  |  |
| --- | --- | --- | --- | --- | --- | --- | --- |
|  | Pus | Urine | SWAB | Sputum | Stool | BAL | ET |
| *Klebsiella pneumoniae* | 7 | 20 | 3 | 2 | 2 | 2 | 3 |
| *Escherichia coli* | 2 | 18 | 4 | 1 | 0 | 0 | 1 |
| *Citrobacter* | 2 | 16 | 0 | 0 | 0 | 0 | 1 |
| *Pseudomonas aeruginosa* | 1 | 6 | 0 | 0 | 0 | 0 | 0 |
| *Acinetobacter* | 2 | 1 | 2 | 1 | 0 | 1 | 0 |
| Total | 14 | 61 | 9 | 4 | 2 | 3 | 4 |

Note# Pus; A thick, whitish-yellow fluid of WBCs (white blood cells), cellular debris, and liquefied tissue. SWAB; a medical device used for the collection of biological samples from the patient body. BAL; Bronchoalveolar lavage, ET; Endotracheal secretion.

### 3.2. Evaluation of antibiotic resistance

To evaluate antibiotic resistance, an antimicrobial susceptibility test (AST) was performed (**Table S1**). The AST of five GNBs, *Klebsiella pneumoniae* (*K*. *pneumoniae*), followed by *Escherichia coli* (*E*. *coli*), *Citrobacter* sp., *Pseudomonas aeruginosa* (*P*. *aeruginosa*), and *Acinetobacter* sp., were determined during our investigation (**Table S1**). Thirty-one antibiotics were used for susceptibility tests to reveal the MDR characteristics of GNBs isolated from patients. All five GNBs (*K*. *pneumoniae*, *E*. *coli*, *Citrobacter* sp., *P*. *aeruginosa*, and *Acinetobacter* sp.) showed the highest (100 %) resistance to ampicillin, ceftazidime, and ertapenem (**Table S1**). *K*. *pneumoniae*, *E*. *coli*, *P*. *aeruginosa*, and *Acinetobacter* sp. were ampicillin/sulbactum-resistant (100 %) (**Table S1**). *K*. *pneumoniae*, *E*. *coli*, *Citrobacter* sp., and *P*. *aeruginosa* were aztreonam-resistant (100 %) (**Table S1**). *K*. *pneumoniae*, *E*. *coli*, *Citrobacter* sp., and *Acinetobacter* sp. were ceftriaxone- and cefuroxime-resistant (100 %) (**Table S1**). *K*. *pneumoniae*, *Citrobacter* sp., and *Acinetobacter* sp. were cefotaxime- and cefixime-resistant (100 %) (**Table S1**). *Citrobacter* sp., *P*. *aeruginosa*, and *Acinetobacter* sp. were cotrimoxazole-, meropenem-, and gentamicin-resistant (100 %) (**Table S1**). *E*. *coli*, *P*. *aeruginosa*, and *Acinetobacter* sp. were ciprofloxacin-resistant (100 %) (**Table S1**). *Citrobacter* sp. and *Acinetobacter* sp. were amikacin-, cefepime-, and tobramycin-resistant (100 %) (**Table S1**). *E*. *coli* and *Citrobacter* sp. were ceftazidime avibactam-resistant (100 %) (**Table S1**). *K*. *pneumoniae* and *E*. *coli* were cefazolin-, norfloxacin-resistant (100 %) (**Table S1**). *K*. *pneumoniae* and *Acinetobacter* sp. were imipenem-, nitrofurantoin-, and piperacillin tazobactam-resistant (100 %) (**Table S1**). *Acinetobacter* sp. was amoxicillin clavulanic acid-, ceftazidime clavulanic acid-, cefoxitin-, and minocycline-resistant (100 %) (**Table S1**). *E*. *coli* was doripenem-resistant (100 %) (**Table S1**). Further, *E*. *coli* was tigecycline- and tetracycline-susceptible (100 %) (**Table S1**).

### 3.3. Identification of MGNB across all the samples

A total of 100 MGNBs belonging to c_Gammaproteobacteria, were identified across all the samples. 28 %, 56 %, and 16 % MGNB were observed in 2021, 2022, and the early part of 2023, respectively (**Fig. 1B**). Further, in the year 2021, 14 adult males, 11 adult females, and 3 children (female) were infected with MGNB, while in the year 2022, 29 adult males, 23 adult females, and 4 children (female) were found to be diseased with MGNB (**Fig. 1C**). Furthermore, in 2023, 10 adult males, 4 adult females, and 2 children (female) were infected with MGNB (**Fig. 1C**). The highest number of adult males, adult female, and children (female) patients with MGNB was revealed in the year 2022 (**Fig. 1C**), while the lowest number of adult male, adult female, and children (female) patients with MGNB were observed in the year 2023 (**Fig. 1C**).

In 100 MGNB isolates, Pseudomonadota was noticed as the dominant phylum among the patient samples. Pseudomonadota alone shared 28, 56, and 16 of the total MGNB isolates in the patient samples collected in 2021, 2022, and 2023, respectively (**Fig. 1B**). The dominant MGNB isolates belonging to Gammaproteobacteria among the patient samples (**Fig. 1D**) depiced the genera *E*. *coli* (28 % isolates), *Citrobacter* sp. (19 % isolates), *K*. *pneumoniae* (40 % isolates), *P*. *aeruginosa* (7 % isolates), and *Acinetobacter* sp. (7 % isolates) (**Fig. S2**).

### 3.4. Specific and shared MGNB population

The number of distinct and shared MGNB isolates present in the patient samples were examined applying a Venn diagram (http://bioinformatics.psb.ugent.be/cgi-bin/liste/Venn/calculate_venn.htpl) (**Fig. 1E**, **Table S2**). Five shared MGNB populations were identified across all samples. *E. coli*, *K*. *pneumoniae*, and *Citrobacter* sp. were shared between three patient samples (2021, 2022, and 2023), while *P*. *aeruginosa* and *Acinetobacter* sp. were shared between only two patient (2021 and 2022) and (2022 and 2023) samples, respectively (**Fig. 1E**, **Table S2**).

### 3.5. Identification of MGNB in patient (with comorbid) samples

*K*. *pneumoniae*, *E*. *coli*, and *Citrobacter* sp. were present in the patient (with comorbid medical conditions) samples collected in 2021 (**Fig. S3A**). *E*. *coli* (in UTI) was highest among the patient (with comorbid) samples (**Fig. S3A**), while *E*. *coli* (in sepsis and hepatic cyst), *K*. *pneumoniae* (in foot gangrene, respiratory disease, and Asthma), *Citrobacter* sp. (in femur fracture) were noted lowest in the patient (with comorbid) samples (**Fig. S3A**).

*K*. *pneumoniae*, *E*. *coli*, *P*. *aeruginosa*, *Citrobacter* sp., and *Acinetobacter* sp. were identified in patient (with comorbid medical conditions) samples collected in 2022 (**Fig. S3B**). *K*. *pneumoniae* (in UTI) was highest among the patient (with comorbid) samples (**Fig. S3B**) while *E*. *coli* (in sepsis, septic shock, cancer, and hemoperitonen), *Citrobacter* sp. (in sepsis, post-CAR, and osteoporosis), *K*. *pneumoniae* (in foot gangrene, respiratory disease, IHD, CKD, hepatic disease, hyperthyroidism) and *P*. *aeruginosa* (in foot gangrene) had the lowest representation in patient (with comorbid) samples (**Fig. S3B**).

*K*. *pneumoniae*, *E*. *coli*, *Citrobacter* sp., and *Acinetobacter* sp. were observed in the patient (with comorbid medical conditions) samples collected in 2023 (**Fig. S3C**). *E*. *coli* (in UTI) was highest among the patient (with comorbid) samples (**Fig. S3C**), while *K*. *pneumoniae* (in sepsis and multiple vessels), *Acinetobacter* sp. (in UTI and in HIE) had the lowest representation in the patient (with comorbid) samples (**Fig. S3C**).

### 3.6. Variance indices

Normal distribution, alpha and beta-diversity (taxonomic variance) of the MGNB population was determined utilizing data from various patient’ samples. The Shapiro-Wilk (S-W) and Anderson-Darling (A-D) normality test were used to determine distributions of MGNB community within the patient samples. (**Table S3**). The S-W analysis for normality varied from 0.800 to 0.888, with a *P-value* of 0.081 to 0.351. The observed A-D normal distribution differed from 0.343 to 0.511, with a *P-value* of 0.098 to 0.315 (**Table S3**). The Alpha (α)- diversity metrics (Shannon, Simpson, Evenness, Brillouin, Fisher alpha, Chao1, and ACE) of the MGNB population were differed notably within the patient samples (**Table 3**). The values of Shannon diversity (1.138 to 1.508), Simpson diversity (0.643 to 0.775), Evenness diversity (0.780 to 1.042), and Brillouin diversity (0.933 to 1.340) (**Table 3**) revealed community structure with moderate species richness and abundance in patient samples. The values of Fisher alpha and Chao1 ranged from 1.277 to 1.712 and 4 to 5, respectively (**Table 3**). The ACE (abundance-based coverage estimator; a species richness index) varied from 4.000 to 5.111, with the increase index in MDR bacteria in 2021 among the patient samples (**Table 3**). Notably, the MDRs in the sample (2022) had more MGNB population than the other samples (**Table 3**). Palaeontological Statistics (PAST, v3) software package was applied to determine the beta (β) diversity of the MGNB population between the patient samples (**Fig. 2A)** collected in 2021, 2022, and 2023. PCoA (principal coordinate analysis, based on the Bray-Kurtis index) was computed using eigenvalues and eigenvectors (coordinates) algorithm from Davis (1986). Before Eigen investigation, PCoA eigen values were produced to the transformation exponent (the power of C), and the definitive index was C = 2. The “Eigen value scaling” measure was applied for each axis, applying the square root of the eigen matrix (value), and the minimum spanning tree preference was based on the picked PCoA matrix. PCoA revealed the existence or absence of MGNB elements between the patient (2021, 2022, and 2023) samples (**Fig. 2A**). Further, analysis of the data, of pair-wise patient sample, comparisons was determined by applying the Bray-Kurtis similarity and dissimilarity index (**Table S4**). Bray-Kurtis pairwise similarity and distance index of the patient samples varied from 0.444 - 1.000 (**Table S4**). Moreover, the Whittaker indexes for the resemblance between MGNB population of the patient samples ranged from 0.11 - 0.25 (**Table S5**).

**Fig 2.**
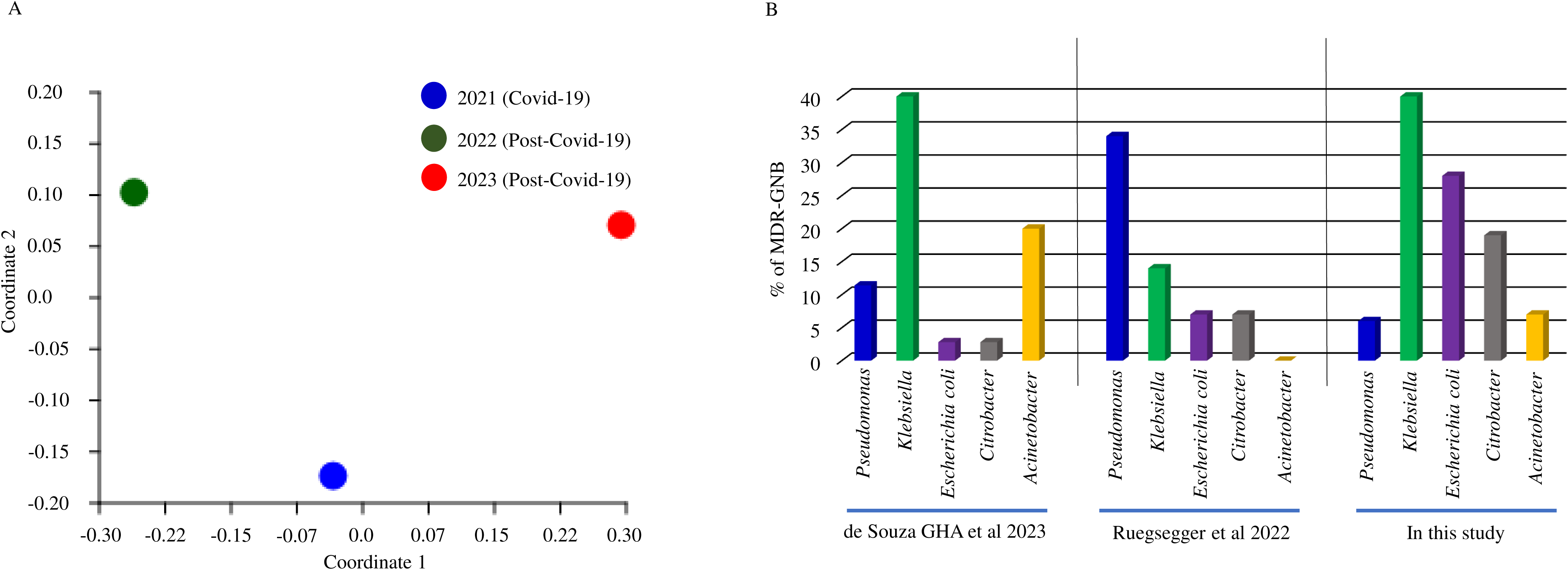
Principal coordinate analysis and comparative overview of MGNB communities identified in patients. (A) Analysis of MDR-gram negative bacterial pathogens diversity in the patient samples collected in different, 2021, 2022, and 2023, years. MDR-gram negative bacterial pathogens diversity in the patient samples using principal coordinate analysis (PCoA-Bray-Kurtis index). (B) Comparative overview of MDR-gram negative bacterial communities identified in patient samples of the Hospital-acquired infection (in this study), de Souza GHA et al (2023), and Ruegsegger et al (2022).

**Table 3.**
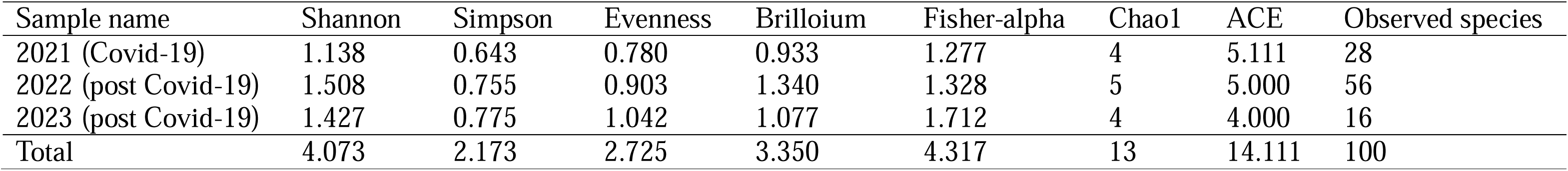
Alpha-diversity metrics of MDR bacterial population in the patient samples.

### 3.7. Comparison of MGNB populations between de Souza GHA et al. (2023), Ruegsegger et al. (2022), and in the present study

In an effort to understand and correlate the MGNB populations of the present study bacterial communities with that present in the study of de Souza GHA et al. (2023), and Ruegsegger et al. (2022) (**Fig. 2B**, **Table 4**). *Pseudomonas* sp. and *Klebsiell* sp. were revealed at 6.0 and 40.0 %, respectively, in the present study, while in the de Souza GHA et al. (2023), they were noted at 11.4 and 40.0 % (**Fig. 2B**, **Table 4**). Further, *E*. *coli* and *Citrobacter* sp. were observed at 28.0 and 19.0 %, respectively in the present study, while in the de Souza GHA et al. (2023) they were noted at 2.8 and 2.8 % (**Fig. 2B**, **Table 4**). Furthermore, *Acinetobacter* sp. was present at 7.0 % in the present study, while in the de Souza GHA et al. (2023) they were revealed at 20.0 % (**Fig. 2B**, **Table 4**).

**Table 4.** Comparative synopsis of MDR bacteria observed in present study patient’ samples (during Hospital stay), de Souza GHA et al., 2023, and Ruegsegger et al., 2022. (ND; Not determined).

| S. No. | MDR microbes | Present study (%) | de Souza GHA et al., 2023, (%) | Ruegsegger et al., 2022, (%) |
| --- | --- | --- | --- | --- |
| 1 | *Pseudomonas* | 6.0 | 11.4 | 34.0 |
| 2 | *Klebsiella* | 40.0 | 40.0 | 14.0 |
| 3 | *Escherichia coli* | 28.0 | 2.8 | 7.0 |
| 4 | *Citrobacter* | 19.0 | 2.8 | 7.0 |
| 5 | *Acinetobacter* | 7.0 | 20.0 | ND |

*Pseudomonas* sp. and *Klebsiella* sp. were present at 6.0 and 40.0 %, respectively, in the present study, while Ruegsegger et al. (2023), reported 34.0 and 14.0 %, respectively (**Fig. 2B**, **Table 4**). Further, *E*. *coli* and *Citrobacter* sp. were present at 28.0 and 19.0 %, respectively, in the present study, while Ruegsegger et al. (2023) reported 7.0 and 7.0 %, respectively (**Fig. 2B**, **Table 4**). Furthermore, *Acinetobacter* sp. was present at 7.0 %, in the present study, while in the Ruegsegger et al. (2023) study, they were not reported (**Fig. 2B**, **Table 4**).

## 4. Discussion

GNB are an important public health issue globally due to their antibiotic resistance characteristic (Oliveira and Reygaert, 2022). The invasion of the host by bacteria and the resulting infection is a versatile mechanism that includes various biological factors, i.e. the host defense mechanism, the prevalence and antibiotic susceptibility pattern of microbial isolates, various bio-physicochemical and genetic attributes (Peterson, 1996). The various virulence factors (persistence, transmissibility (ratio of output to input), cling to host cells, host cells invasion, toxigenicity, and the capability to elude or live the host’s defence mechanism) of bacteria have been extensively investigated and have shown to have a negative impact on patient’s health, particularly when a patient is immunocompromised due to severe disease (Freeman et al., 2020; Moradi et al., 2021). Moreover, it would be valuable to investigate whether patient genotypes can influence survival by influencing the number of MGNB within them (increased or decreased count of MGNB).

ICU-MDR percentages are maximum than in other hospital wards (**Table 1**) due to the multiple associations between the patient’s underlying medical conditions, hospital ward type, LOS (length of stay), and employment of various invasive tools (Erbay et al., 2003; Inweregbu et al., 2005; Vincent et al., 2009).

Among the 100 patients investigated, 85 (85 %) were discharged, while 5 (5 %) expired in the hospital (**Fig. 1A**). An overall lower mortality rate was found among elderly, severe patients (with comorbidities) admitted to the different units of the hospital. Our outcomes are not in concurrence with the investigations of Juliana et al. (2022) and Boorgula et al. (2022), who revealed a higher death rate in serious patients. Taking this observation into consideration, we speculate that the lower mortality rate, in our study, might be due to the higher immunity of studied patients during their hospitalization (Parohan et al., 2020; Weiss and Murdoch, 2020; Ejaz et al., 2020). However, such intrinsic protective factors have yet to be determined in specific populations to make the above claims regarding mortality.

In our investigation, *K*. *pneumoniae*, *E*. *coli*, *Citrobacter* sp., *Acinetobacter* sp., *P*. *aeruginosa* were the most frequent isolates (GNB) in various patient’ samples (Vijay et al., 2021; Sharifipour et al., 2020). GNB isolates were higher in adult males than in adult females and children (female) (**Fig. 1C**). With this observation in view, we hypothesize that a higher level of an enzyme called carboxypeptidase (ACE 2, angiotensin-converting enzyme 2) in men and the predisposition of adult females (at reproductive age) to autoimmune disorder than infectious diseases might be the two reasons for higher MDR-GNB male than in female. Lifestyles of men (heavy smoking and drinking) make them more susceptible to infections (Ramírez-Soto et al., 2021; Bwire, 2020). Nevertheless, this is yet to be elucidated.

Regarding AST, we revealed *Acinetobacter* sp. to be the most (100 %) resistant strain when measured against twenty antibiotics (**Table S1**), while *Klebsiella* sp., was found to be 100 % resistant against eleven antibiotics (**Table S1**). All these five different bacterial species (**Table S1**) were resistant to numerous antimicrobial agents, which gives them the title of multidrug-resistant bacteria (Giannitsioti et al., 2022). Surprisingly antibacterial resistance, in this study, is related to a decrease in the mortality rate compared with the previous study (Tanwar et al., 2014).

Our data provides evidence of an increase in antimicrobial resistance post-Covid-19 (**Fig. 1C**, data collected in 2022). This is in keeping with previous reports that have mentioned a similar pattern of increased antimicrobial resistance in specific locations (Shomuyiwa et al., 2022). Indiscriminate and irrational antimicrobial use and weak antimicrobials regulatory ecosystem, which could be partially attributed to health systems disruption by inadequate access to health services during the Covid-19 pandemic, are suggested causes for the rapid growth of antimicrobial resistance in the community (Lobie et al., 2021; Shomuyiwa et al., 2022). Given the rapid increase in antimicrobial resistance post-Covid-19 in our study, it is important to further analyze how the policies adopted to manage Covid-19 (i.e. widespread antibiotic and disinfectant usage) affect or have a long-lasting consequence on antimicrobial resistance (AMR) (Nieuwlaat et al., 2021). Furthermore, programs at a governmental level need to be adopted widely, particularly in low- and middle-income countries (LMIC), to contain the possible increase in AMR (Lucien et al., 2021).

## 5. Conclusion

In conclusion, outcomes achieved from the present investigation revealed that MDR- microbes present in patients are predominantly elements of the gamma-proteobacteria. Antibacterial resistance has an important impact on therapeutics and the outcome of bacterial infection. In this study, five gram-negative bacteria, *K*. *pneumoniae*, *E*. *coli*, *Citrobacter* sp., *Acinetobacter* sp., and *P*. *aeruginosa*, had increased resistance to the antibacterial agents applied. Therefore, it is essential that the public health and drug susceptibility patterns of microorganisms initiating pathogen infection are continually observed to instruct clinical trials and treatment strategies, thereby reducing the appearance of multi-drug-resistant microbial pathogens.

## Data Availability

All data produced in the present work are contained in the manuscript

## Declarations

### Ethics approval and consent to participate

Patient samples and data were collected from the Symbiosis University Hospital and Research Centre (SUHRC), the Symbiosis International University, Pune, Maharashtra, India.

### Consent to publish

Not applicable.

### Availability of data and materials

All data is given in the main body of the manuscript; materials are available from the authors. All sequence data is added to manuscript as supplementary files (named as Figure S1 to S3, and Tables S1 to S6).

### Authors’ contributions

A.O. made substantial contributions to conception, was involved in analysis & interpretation of data, writing, revising and drafting the manuscript, and given final approval of the version to be published. D.W. made substantial contributions to writing discussion, editing the manuscript, was involved in revising the manuscript, and given final approval of the version to be published. S.K. made substantial contributions to collect the data from SUHRC and given final approval of the version to be published. All authors reviewed the manuscript and given final approval of the version to be published.

### Authors’ Information

Symbiosis Institute of Health Sciences, Symbiosis International (Deemed University), Lavale (Hill Base), Pune 412115, Maharashtra, India.

## Acknowledgments

We thank director, Symbiosis Institute of Health Sciences (SIHS) and Head, Symbiosis University Hospital and Research Centre (SUHRC), Pune for kind support.

## Supplementary material

**Fig. S1**. Overview of MDR-gram negative bacterial pathogens collected from the gender wise patients.

**Fig. S2.** Relative abundance of different MDR-gram negative bacterial pathogens among all patient samples at the level of species.

**Fig. S3**. Description of MDR-gram negative bacterial pathogens among the patient with comorbid condition collected in different, (A) 2021, (B) 2022, and (C) 2023, years from hospital.

**Table S1**. Antimicrobial susceptibility tests. Antimicrobial susceptibility tests of MGNB pathogens collected from patient samples.

**Table S2**. Labeling of specific and shared MGNB communities among the patient samples during Covid-19 and post-Covid-19 year. (A) Specific MGNB communities (specific and shared) in patient samples. (B) Shared MGNB communities in patient samples.

**Table S3** Statistics of MGNB collected different years from the patient samples.

**Table S4**. Bray-Kurtis pairwise similarity and distance (dissimilarity) index of MGNB population among the patient samples collected in 2021, 2022, and 2023.

**Table S5** Beta-diversity, Whittaker indexes, and pair-wise comparisons between the patient samples based on MGNB populations.

