## Supplementary Figure S1 for "A comparative study of multidrug resistant bacteria (MDRB) isolated from various patients in different wards of a hospital during post Covid-19"

### Slide 1
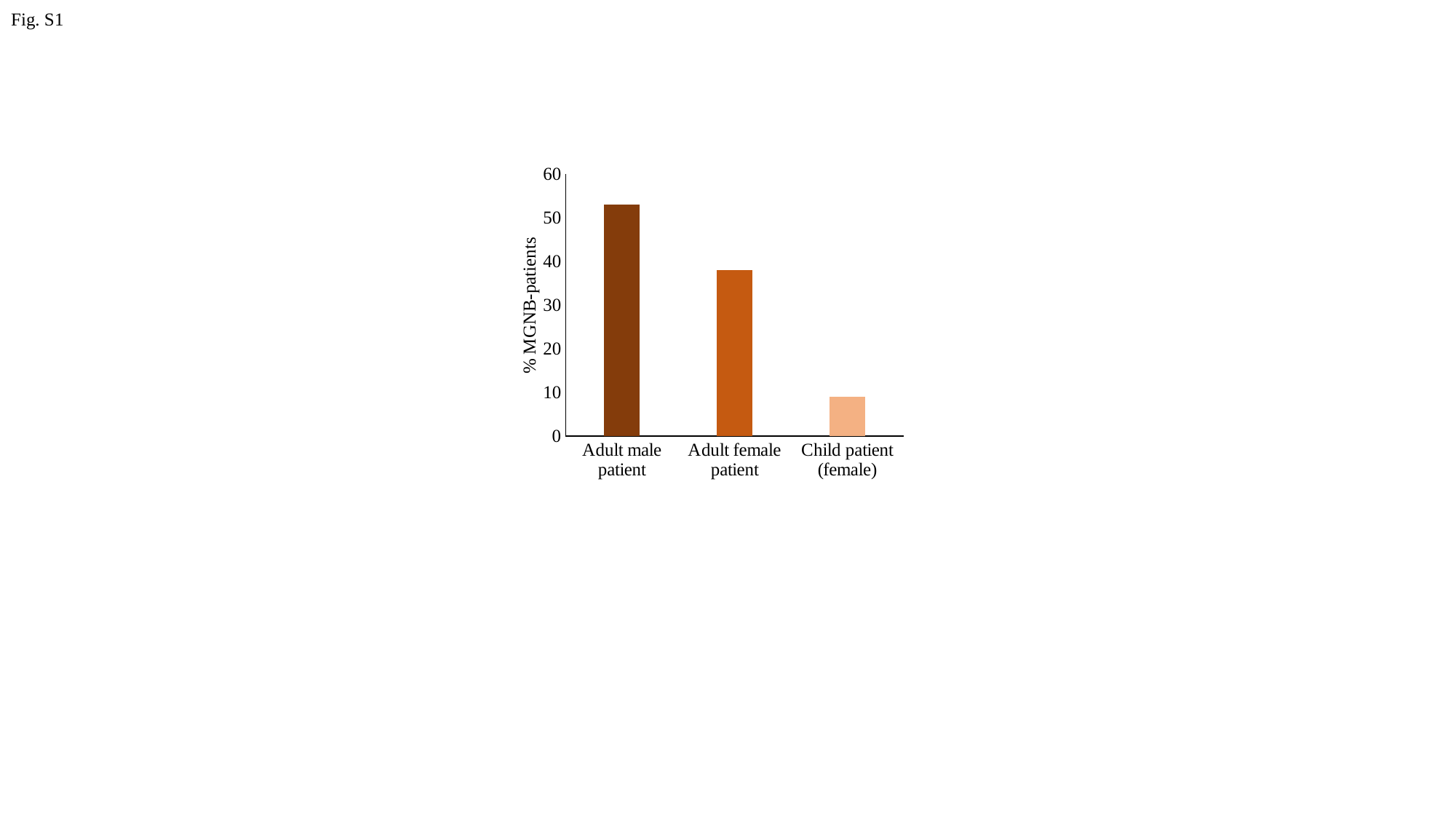

Fig. S1
#### Chart
| Category | |
|---|---|
| Adult male patient | 53.0 |
| Adult female patient | 38.0 |
| Child patient (female) | 9.0 |% MGNB-patients
