## Supplementary Figure S2 for "A comparative study of multidrug resistant bacteria (MDRB) isolated from various patients in different wards of a hospital during post Covid-19"

### Slide 1
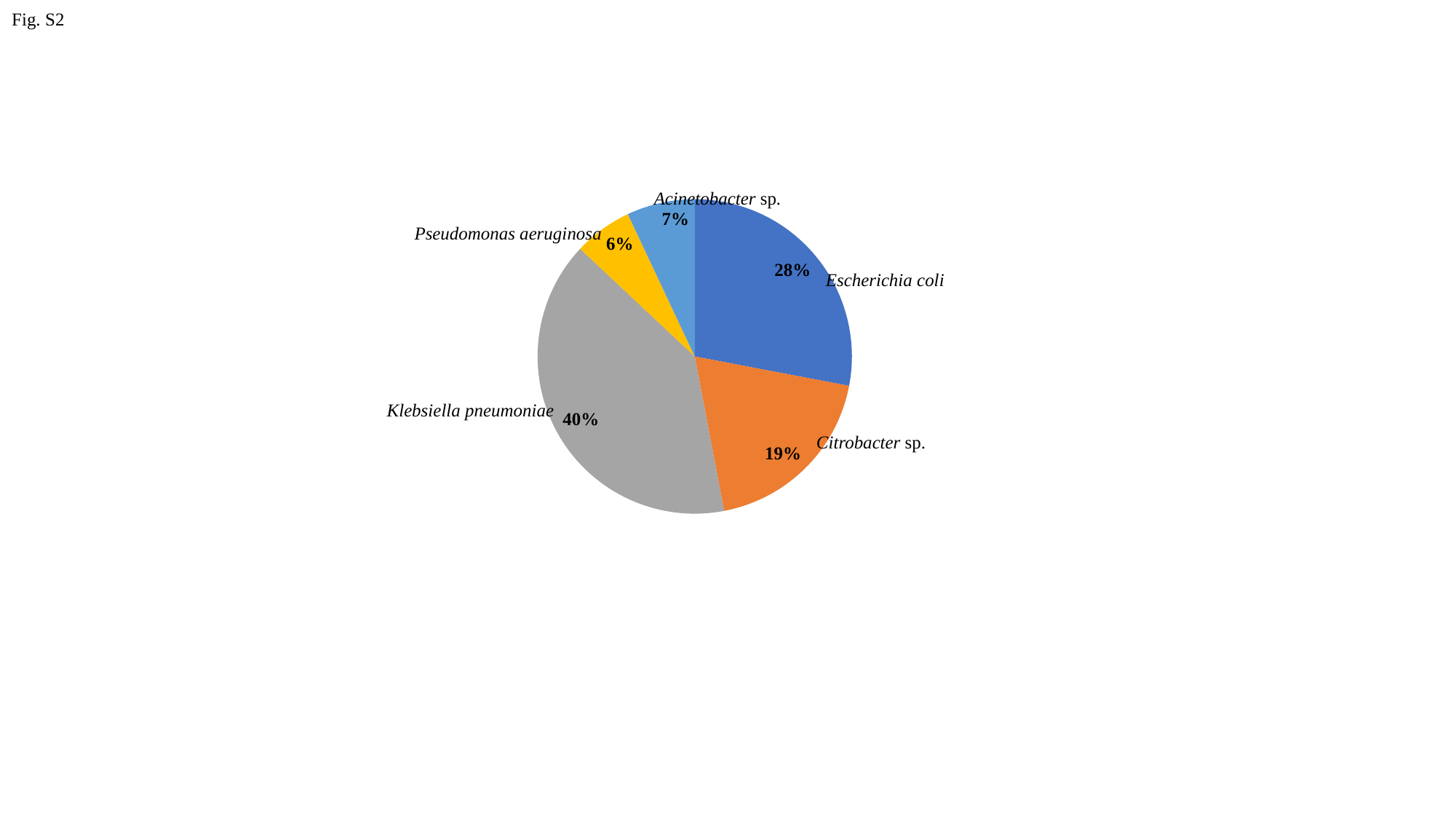

Fig. S2
Acinetobacter sp.
#### Chart
| Category | |
|---|---|
| Eschirechia coli | 28.0 |
| Citrobacter | 19.0 |
| Kliebsiella pneumoniae | 40.0 |
| Pseudomonas aeruginosa | 6.0 |
| Acinetobacter | 7.0 |Pseudomonas aeruginosa
Escherichia coli
Klebsiella pneumoniae
Citrobacter sp.
