## Supplementary Figure S3 for "A comparative study of multidrug resistant bacteria (MDRB) isolated from various patients in different wards of a hospital during post Covid-19"

### Slide 1
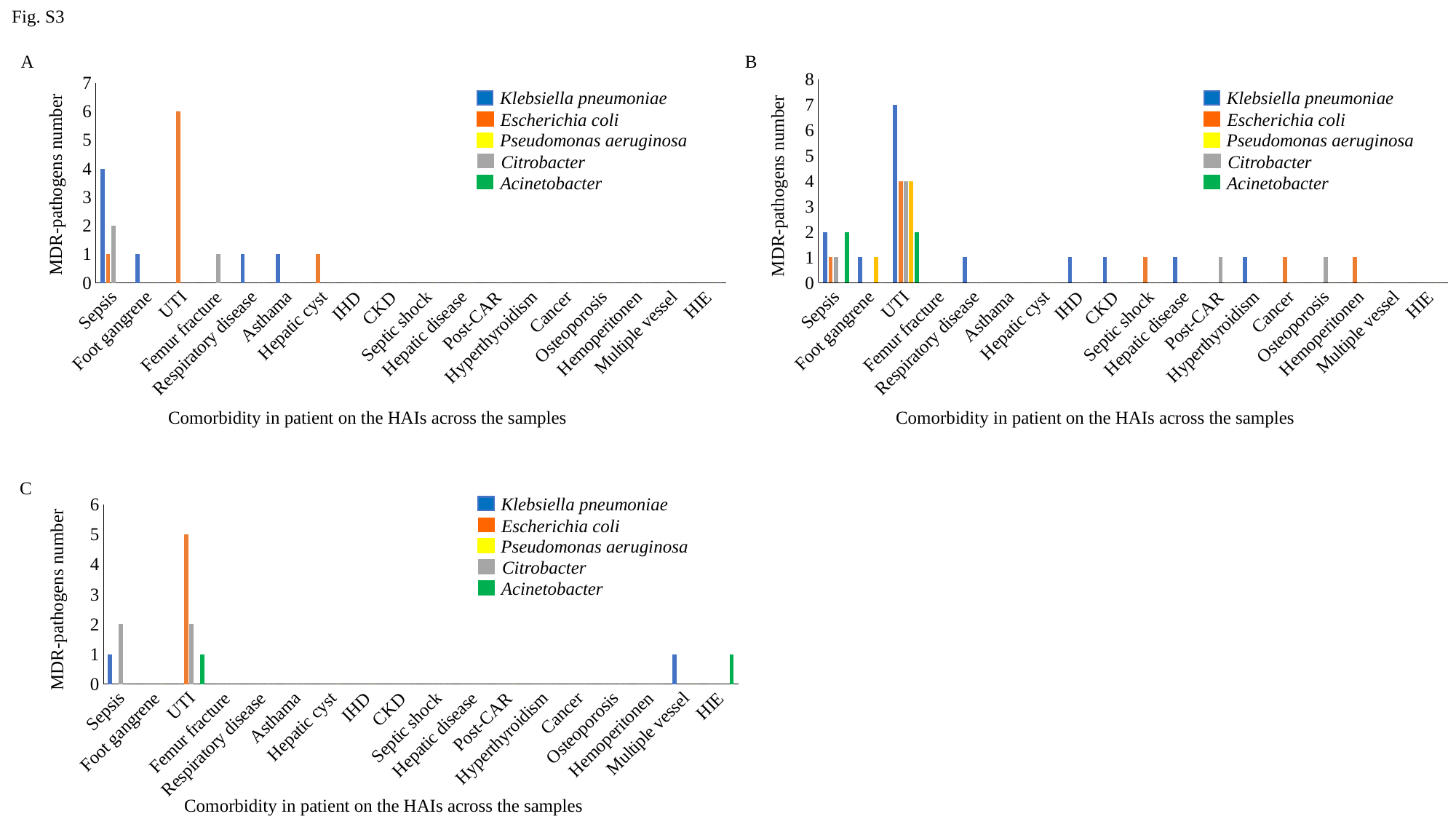

Fig. S3
A
#### Chart
| Category | Kliebsiella pneumoniae | Escherichia coli | Citrobacter | Pseudomonas aeruginosa | Acinetobacter |
|---|---|---|---|---|---|
| Sepsis | 4.0 | 1.0 | 2.0 | 0.0 | 0.0 |
| Foot gangrene | 1.0 | 0.0 | 0.0 | 0.0 | 0.0 |
| UTI | 0.0 | 6.0 | 0.0 | 0.0 | 0.0 |
| Femur fracture | 0.0 | 0.0 | 1.0 | 0.0 | 0.0 |
| Respiratory disease | 1.0 | 0.0 | 0.0 | 0.0 | 0.0 |
| Asthama | 1.0 | 0.0 | 0.0 | 0.0 | 0.0 |
| Hepatic cyst | 0.0 | 1.0 | 0.0 | 0.0 | 0.0 |
| IHD | 0.0 | 0.0 | 0.0 | 0.0 | 0.0 |
| CKD | 0.0 | 0.0 | 0.0 | 0.0 | 0.0 |
| Septic shock | 0.0 | 0.0 | 0.0 | 0.0 | 0.0 |
| Hepatic disease | 0.0 | 0.0 | 0.0 | 0.0 | 0.0 |
| Post-CAR | 0.0 | 0.0 | 0.0 | 0.0 | 0.0 |
| Hyperthyroidism | 0.0 | 0.0 | 0.0 | 0.0 | 0.0 |
| Cancer | 0.0 | 0.0 | 0.0 | 0.0 | 0.0 |
| Osteoporosis | 0.0 | 0.0 | 0.0 | 0.0 | 0.0 |
| Hemoperitonen | 0.0 | 0.0 | 0.0 | 0.0 | 0.0 |
| Multiple vessel | 0.0 | 0.0 | 0.0 | 0.0 | 0.0 |
| HIE | 0.0 | 0.0 | 0.0 | 0.0 | 0.0 |MDR-pathogens number
Comorbidity in patient on the HAIs across the samples
Klebsiella pneumoniae
Escherichia coli
Pseudomonas aeruginosa
Citrobacter
Acinetobacter
B
#### Chart
| Category | Kliebsiella pneumoniae | Escherichia coli | Citrobacter | Pseudomonas aeruginosa | Acinetobacter |
|---|---|---|---|---|---|
| Sepsis | 2.0 | 1.0 | 1.0 | 0.0 | 2.0 |
| Foot gangrene | 1.0 | 0.0 | 0.0 | 1.0 | 0.0 |
| UTI | 7.0 | 4.0 | 4.0 | 4.0 | 2.0 |
| Femur fracture | 0.0 | 0.0 | 0.0 | 0.0 | 0.0 |
| Respiratory disease | 1.0 | 0.0 | 0.0 | 0.0 | 0.0 |
| Asthama | 0.0 | 0.0 | 0.0 | 0.0 | 0.0 |
| Hepatic cyst | 0.0 | 0.0 | 0.0 | 0.0 | 0.0 |
| IHD | 1.0 | 0.0 | 0.0 | 0.0 | 0.0 |
| CKD | 1.0 | 0.0 | 0.0 | 0.0 | 0.0 |
| Septic shock | 0.0 | 1.0 | 0.0 | 0.0 | 0.0 |
| Hepatic disease | 1.0 | 0.0 | 0.0 | 0.0 | 0.0 |
| Post-CAR | 0.0 | 0.0 | 1.0 | 0.0 | 0.0 |
| Hyperthyroidism | 1.0 | 0.0 | 0.0 | 0.0 | 0.0 |
| Cancer | 0.0 | 1.0 | 0.0 | 0.0 | 0.0 |
| Osteoporosis | 0.0 | 0.0 | 1.0 | 0.0 | 0.0 |
| Hemoperitonen | 0.0 | 1.0 | 0.0 | 0.0 | 0.0 |
| Multiple vessel | 0.0 | 0.0 | 0.0 | 0.0 | 0.0 |
| HIE | 0.0 | 0.0 | 0.0 | 0.0 | 0.0 |MDR-pathogens number
Comorbidity in patient on the HAIs across the samples
Klebsiella pneumoniae
Escherichia coli
Pseudomonas aeruginosa
Citrobacter
Acinetobacter
C
#### Chart
| Category | Kliebsiella pneumoniae | Escherichia coli | Citrobacter | Pseudomonas aeruginosa | Acinetobacter |
|---|---|---|---|---|---|
| Sepsis | 1.0 | 0.0 | 2.0 | 0.0 | 0.0 |
| Foot gangrene | 0.0 | 0.0 | 0.0 | 0.0 | 0.0 |
| UTI | 0.0 | 5.0 | 2.0 | 0.0 | 1.0 |
| Femur fracture | 0.0 | 0.0 | 0.0 | 0.0 | 0.0 |
| Respiratory disease | 0.0 | 0.0 | 0.0 | 0.0 | 0.0 |
| Asthama | 0.0 | 0.0 | 0.0 | 0.0 | 0.0 |
| Hepatic cyst | 0.0 | 0.0 | 0.0 | 0.0 | 0.0 |
| IHD | 0.0 | 0.0 | 0.0 | 0.0 | 0.0 |
| CKD | 0.0 | 0.0 | 0.0 | 0.0 | 0.0 |
| Septic shock | 0.0 | 0.0 | 0.0 | 0.0 | 0.0 |
| Hepatic disease | 0.0 | 0.0 | 0.0 | 0.0 | 0.0 |
| Post-CAR | 0.0 | 0.0 | 0.0 | 0.0 | 0.0 |
| Hyperthyroidism | 0.0 | 0.0 | 0.0 | 0.0 | 0.0 |
| Cancer | 0.0 | 0.0 | 0.0 | 0.0 | 0.0 |
| Osteoporosis | 0.0 | 0.0 | 0.0 | 0.0 | 0.0 |
| Hemoperitonen | 0.0 | 0.0 | 0.0 | 0.0 | 0.0 |
| Multiple vessel | 1.0 | 0.0 | 0.0 | 0.0 | 0.0 |
| HIE | 0.0 | 0.0 | 0.0 | 0.0 | 1.0 |MDR-pathogens number
Comorbidity in patient on the HAIs across the samples
Klebsiella pneumoniae
Escherichia coli
Pseudomonas aeruginosa
Citrobacter
Acinetobacter
