## Supplementary Table S1 for "A comparative study of multidrug resistant bacteria (MDRB) isolated from various patients in different wards of a hospital during post Covid-19"

Table S1׀ An antibiotic sensitivity test of MDR bacteria isolated from patient samples (2021-2023).

|  | | GNB names and numbers (%) | | | | | | | |
| --- | --- | --- | --- | --- | --- | --- | --- | --- | --- |
| S. No. | Antibiotics | | Disk concentration  (mcg) | Interpretation  (S, R, I) | *Klebsiella* spp.  n=38 | *Escherichia coli*  n=28 | *Citrobacter* spp.  n=19 | *Pseudomonas* spp.  n=6 | *Acinetobacter*  n=7 |
| 1 | Ampicillin-sulbactam | | 10/10 | S  R  I | 0 (0)  38 (100 %)  0 (0) | 0 (0)  28 (100 %)  0 (0) | 2 (10.5 %)  17 (89.5 %)  0 (0) | 0 (0)  6 (100 %)  0 (0) | 0 (0)  7 (100 %)  0 (0) |
| 2 | Amoxicillin-clavulanate | | 20/10 | S  R  I | 7 (18.4 %)  31 (81.6 %)  0 (0) | 3 (10.7 %)  25 (90 %)  0 (0) | 1 (5.3 %)  18 (94.7 %)  0 (0) | 0 (0)  0 (0)  0 (0) | 0 (0)  7 (100 %)  0 (0) |
| 3 | Aztreonam | | 30 | S  R  I | 0 (0)  38 (100 %)  0 (0) | 0 (0)  28 (100 %)  0 (0) | 0 (0)  19 (100 %)  0 (0) | 0 (0)  6 (100 %)  0 (0) | 0 (0)  0 (0)  0 (0) |
| 4 | Ampicillin | | 10 | S  R  I | 0 (0)  38 (100 %)  0 (0) | 0 (0)  28 (100 %)  0 (0) | 0 (0)  19 (100 %)  0 (0) | 0 (0)  6 (100 %)  0 (0) | 0 (0)  7 (100 %)  0 (0) |
| 5 | Amikacin | | 30 | S  R  I | 6 (16.8 %)  32 (84.2 %)  0 (0) | 21 (75 %)  7 (25 %)  0 (0) | 0 (0)  19 (100 %)  0 (0) | 0 (0)  6 (100 %)  0 (0) | 0 (0)  7 (100 %)  0 (0) |
| 6 | Cefotaxime | | 30 | S  R  I | 0 (0)  38 (100 %)  0 (0) | 0 (0)  27 (96.4 %)  1 (3.6 %) | 0 (0)  19 (100 %)  0 (0) | -  -  - | 0 (0)  7 (100 %)  0 (0) |
| 7 | Ceftazidime-clavulanic acid | | 30/10 | S  R  I | 7 (18.4 %)  31 (81.6 %)  0 (0) | 4 (14.3 %)  23 (82.1 %)  1 (3.6 %) | 1 (5.3 %)  18 (94.7 %)  0 (0) | -  -  - | 0 (0)  7 (100 %)  0 (0) |
| 8 | Tigecycline | | 16 | S  R  I | -  -  - | 28 (100 %)  0 (0)  0 (0) | 18 (94.7 %)  0 (0)  1 (5.3 %) | -  -  - | 0 (0)  0 (0)  0 (0) |
| 9 | Ceftazidime-avibactam | | 30/20 | S  R  I | 6 (15.8 %)  32 (84.2 %)  0 (0) | 0 (0)  28 (100 %)  0 (0) | 0 (0)  19 (100 %)  0 (0) | -  -  - | 0 (0)  0 (0)  0 (0) |
| 10 | Fosfomycin | | 200 | S  R  I | 19 (50 %)  19 (50 %)  0 (0) | 27 (96.4 %)  1 (3.6 %)  0 (0) | 0 (0)  0 (0)  0 (0) | -  -  - | 0 (0)  0 (0)  0 (0) |
| 11 | Tobramycin | | 10 | S  R  I | 5 (13.2 %)  33 (86.8 %)  0 (0) | 6 (21.4 %)  22 (78.6 %)  0 (0) | 0 (0)  19 (100 %)  0 (0) | -  -  - | 0 (0)  7 (100 %)  0 (0) |
| 12 | Ceftriaxone | | 30 | S  R  I | 0 (0)  38 (100 %)  0 (0) | 0 (0)  28 (100 %)  0 (0) | 0 (0)  19 (100 %)  0 (0) | -  -  - | 0 (0)  7 (100 %)  0 (0) |
| 13 | Cefuroxime | | 30 | S  R  I | 0 (0)  38 (100 %)  0 (0) | 0 (0)  28 (100 %)  0 (0) | 0 (0)  19 (100 %)  0 (0) | -  -  - | 0 (0)  7 (100 %)  0 (0) |
| 14 | Cefazolin | | 30 | S  R  I | 0 (0)  38 (100 %)  0 (0) | 0 (0)  28 (100 %)  0 (0) | 0 (0)  0 (0)  0 (0) | -  -  - | 0 (0)  0 (0)  0 (0) |
| 15 | Norfloxacin | | 10 | S  R  I | 0 (0)  38 (100 %)  0 (0) | 0 (0)  28 (100 %)  0 (0) | 0 (0)  0 (0)  0 (0) | -  -  - | 0 (0)  0 (0)  0 (0) |
| 16 | Cefoxitin | | 34 | S  R  I | 5 (13.2 %)  33 (86.8 %)  0 (0) | 3 (10.7 %)  24 (85.7 %)  1 (4) | 1 (5.3 %)  18 (96.7 %)  0 (0) | -  -  - | 0 (0)  7 (100 %)  0 (0) |
| 17 | Ceftazidime | | 30 | S  R  I | 0 (0)  38 (100 %)  0 (0) | 0 (0)  28 (100 %)  0 (0) | 0 (0)  19 (100 %)  0 (0) | 0 (0)  6 (100 %)  0 (0) | 0 (0)  7 (100 %)  0 (0) |
| 18 | Cefepime | | 30 | S  R  I | 6 (15.8 %)  32 (84.2 %)  0 (0) | 4 (14.3 %)  24 (85.7 %)  0 (0) | 0 (0)  19 (100 %)  0 (0) | 0 (0)  0 (0)  0 (0) | 0 (0)  7 (100 %)  0 (0) |
| 19 | Ciprofloxacin | | 5 | S  R  I | 3 (7.9 %)  35 (92.1 %)  0 (0) | 0 (0)  28 (100 %)  0 (0) | 1 (5.3 %)  18 (96.7 %)  0 (0) | 0 (0)  6 (100 %)  0 (0) | 0 (0)  7 (100 %)  0 (0) |
| 20 | Cotrimoxazole | | 1.25/23.75 | S  R  I | 6 (15.8 %)  32 (84.2 %)  0 (0) | 1 (3.6 %)  27 (96.4 %)  0 (0) | 0 (0)  19 (100 %)  0 (0) | 0 (0)  6 (100 %)  0 (0) | 0 (0)  7 (100 %)  0 (0) |
| 21 | Imipenem | | 10 | S  R  I | 14 (36.8 %)  24 (63.2 %)  0 (0) | 6 (21.4 %)  21 (75 %)  1 (3.6 %) | 1 (5.3 %)  18 (94.7 %)  0 (0) | 0 (0)  6 (100 %)  0 (0) | 0 (0)  7 (100 %)  0 (0) |
| 22 | Minocycline | | 30 | S  R  I | 15 (39.2 %)  15 (39.2 %)  8 (21.1 %) | 3 (10.7 %)  13 (46.4 %)  12 (44 %) | 0 (0)  0 (0)  19 (100 %) | 0 (0)  0 (0)  0 (0) | 0 (0)  7 (100 %)  0 (0) |
| 23 | Meropenem | | 10 | S  R  I | 6 (15.8 %)  32 (84.2 %)  0 (0) | 1 (3.6 %)  26 (92.8 %)  1 (3.6 %) | 0 (0)  19 (100 %)  0 (0) | 0 (0)  6 (100 %)  0 (0) | 0 (0)  7 (100 %)  0 (0) |
| 24 | Levofloxacin | | 5 | S  R  I | 4 (10.5 %)  34 (89.5 %)  0 (0) | 0 (0)  28 (100 %)  0 (0) | 2 (10.5 %)  17 (89.5 %)  0 (0) | 0 (0)  6 (100 %)  0 (0) | 0 (0)  7 (100 %)  0 (0) |
| 25 | Nitrofurantoin | | 300 | S  R  I | 11 (29 %)  27 (71 %)  0 (0) | 19 (67.8 %)  8 (28.6 %)  1 (3.6 %) | 1 (5.3 %)  17 (89.5 %)  1 (5.3 %) | 0 (0)  6 (100 %)  0 (0) | 0 (0)  7 (100 %)  0 (0) |
| 26 | Piperacillin-tazobactam | | 100/10 | S  R  I | 7 (18.4 %)  28 (73.6 %)  3 (8 %) | 6 (21.4 %)  22 (78.6 %)  0 (0) | 2 (10.5 %)  17 (89.5 %)  0 (0) | 0 (0)  6 (100 %)  0 (0) | 0 (0)  7 (100 %)  0 (0) |
| 27 | Ertapenem | | 10 | S  R  I | 0 (0)  38 (100 %)  0 (0) | 0 (0)  28 (100 %)  0 (0) | 0 (0)  19 (100 %)  0 (0) | 0 (0)  6 (100 %)  0 (0) | 0 (0)  7 (100 %)  0 (0) |
| 28 | Gentamicin | | 10 | S  R  I | 4 (10.5 %)  34 (89.5 %)  0 (0) | 5 (17.9 %)  23 (82.1 %)  0 (0) | 0 (0)  19 (100 %)  0 (0) | 0 (0)  6 (100 %)  0 (0) | 0 (0)  7 (100 %)  0 (0) |
| 29 | Cefixime | | 5 | S  R  I | 0 (0)  38 (100 %)  0 (0) | 0 (0)  0 (0)  0 (0) | 0 (0)  19 (100 %)  0 (0) | 0 (0)  0 (0)  0 (0) | 0 (0)  7 (100 %)  0 (0) |
| 30 | Tetracycline | | 30 | S  R  I | -  -  - | 28 (100 %)  0 (0)  0 (0) | 0 (0)  0 (0)  0 (0) | 0 (0)  0 (0)  0 (0) | 0 (0)  0 (0)  0 (0) |
| 31 | Doripenem | | 10 | S  R  I | -  -  - | 0 (0)  28 (100 %)  0 (0) | 0 (0)  0 (0)  0 (0) | 0 (0)  0 (0)  0 (0) | 0 (0)  0 (0)  0 (0) |

Clinical and laboratory standards institute (CLSI) guidelines (2022) applied for analysing antimicrobial susceptibility test outcomes.

Note: S; Sensitive, I; Intermediate, R; Resistance.
