## Supplementary Table S2 for "A comparative study of multidrug resistant bacteria (MDRB) isolated from various patients in different wards of a hospital during post Covid-19"

Table S2׀ Labeling of specific and shared MDR-gram negative bacterial pathogens among the patient samples, 2021-2023 (Covid-19/ or post Covid-19) period.

A

| Year (Samples collected) | Number of bacteria | Number of unique bacteria |
| --- | --- | --- |
| 2021 (Covid-19) | 28 | 4 |
| 2022 (post Covid-19) | 56 | 5 |
| 2023 (post Covid-19) | 16 | 4 |
| Overall number of unique MDR-pathogens | | 5 |

B

| Year (Samples collected) | Total bacteria | Bacteria name |
| --- | --- | --- |
| 2021 2022 2023 | 3 | *Escherichia coli Klebsiella pneumoniae Citrobacter* |
| 2021 2022 | 1 | *Pseudomonas aeruginosa* |
| 2022 2023 | 1 | *Acinetobacter* |
