## Supplementary Table S3 for "A comparative study of multidrug resistant bacteria (MDRB) isolated from various patients in different wards of a hospital during post Covid-19"

Table S3׀ Statistics and normal distributions of bacterial isolates from the patient’ samples collected during, Covid-19 or post Covid-19 period, hospital stay in various years (2021-2023).

A

Statistics of bacterial isolates from the patient’ samples.

|  | 2021 (Covid-19) | 2022 (post Covid-19) | 2023 (post Covid-19) |
| --- | --- | --- | --- |
| N | 5 | 5 | 5 |
| Min | 0 | 5 | 0 |
| Max | 12 | 22 | 5 |
| Sum | 28 | 56 | 16 |
| Mean | 5.6 | 11.2 | 3.2 |
| Standard deviation (±) | 5.94 | 6.76 | 2.16 |
| Standard error (±) | 2.65 | 3.02 | 0.97 |

B

Normal distributions of bacterial isolates from the patient’ samples.

|  | 2021 (Covid-19) | 2022 (post Covid-19) | 2023 (post Covid-19) |
| --- | --- | --- | --- |
| N | 5 | 5 | 5 |
| Shapiro-Wilk W | 0.80 | 0.89 | 0.87 |
| P (normal) | 0.08 | 0.35 | 0.27 |
| Anderson-Darling A | 0.51 | 0.34 | 0.36 |
| P (normal) | 0.09 | 0.31 | 0.28 |
