## Supplementary Table S4 for "A comparative study of multidrug resistant bacteria (MDRB) isolated from various patients in different wards of a hospital during post Covid-19"

Table S4׀ Bray-Kurtis pair-wise comparisons, similarity and distance indices, between the patient samples based on the bacterial populations. The patient samples (during hospital stay); 2021 (Covid-19), 2022 (post-Covid-19), and 2023 (post-Covid-19).

|  | 2021 (Covid-19) | 2022 (post Covid-19) | 2023 (post Covid-19) |
| --- | --- | --- | --- |
| 2021 (Covid-19) | 1 | 0.6428 | 0.5909 |
| 2022 (post Covid-19) | 0.6428 | 1 | 0.4444 |
| 2023 (post Covid-19) | 0.5909 | 0.4444 | 1 |
