## Supplementary Table S5 for "A comparative study of multidrug resistant bacteria (MDRB) isolated from various patients in different wards of a hospital during post Covid-19"

Table S5׀ Beta-diversity, Whittaker indexes, and pair-wise comparisons between the patient samples based on MGNB populations.

|  | 2021 (Covid-19) | 2022 (post Covid-19) | 2023 (post Covid-19) |
| --- | --- | --- | --- |
| 2021 (Covid-19) | 0 | 0.111 | 0.25 |
| 2022 (post Covid-19) | 0.111 | 0 | 0.111 |
| 2023 (post Covid-19) | 0.25 | 0.111 | 0 |
